# Vaccine effectiveness of BNT162b2 mRNA Covid-19 Vaccine in Children below 5 Years in German Primary Care

**DOI:** 10.1101/2023.05.05.23289209

**Authors:** Christoph Strumann, Otavio Ranzani, Jeanne Moor, Reinhard Berner, Nicole Töpfner, Cho-Ming Chao, Matthias B. Moor

## Abstract

**Background:** Despite the approval of BNT162b2 mRNA vaccine for children aged 6 months to 4 years by the European Medicines Agency (EMA) and the Federal Drug Administration (FDA) in 2022, no data on vaccine effectiveness (VE) of BNT162b2 are available in this age group. We here report on the VE of BNT162b2 during an Omicron BA.1-2 dominant period.

**Methods:** An authentication-based retrospective survey was performed between April 14th 2022 and May 9th 2022 in individuals that had registered children for off-label SARS-CoV-2 vaccination in Germany. We used Cox regression to estimate relative VE of two BNT162b2 doses, with the period between first and second vaccine dose as reference period (24.8+-0.6 days) and >=7 days after Dose 2 to before Dose 3 as post-vaccination period (59.5+-23.6 days).

**Results:** The present analysis included 4615 children aged 2.8+-1.2 years (mean+-standard deviation) who had received their first dose of BNT162b2 on January 1st 2022 or thereafter. VE was substantial for protection from any SARS-CoV-2 infection (VE: 53.1% [95% confidence interval (CI): 36.3-69.6%], p<0.001), symptomatic SARS-CoV-2 infections (VE: 57.5% [95% CI: 40.8-74.2%], p<0.001), and SARS-CoV-2 infections leading to medication use (VE: 66.2% [95% CI: 43.7-88.7%], p<0.001). Differences in dosage of BNT162b2 yielded no change in VE.

**Conclusion:** This study offers a first industry-independent insight in the potential VE of two doses of the BNT162b2 vaccine in children aged below 5 years, as currently only immunogenicity data by the manufacturer Pfizer/BioNTech are available. Limitations include the retrospective study design, and that the reported VE does not necessarily correspond to currently circulating SARS-CoV-2 variants.

## Main

Despite the approval of BNT162b2 mRNA vaccine (Pfizer/BioNTech vaccine Comirnaty®) for children aged 6 months to 4 years by the European Medicines Agency (EMA) and the Federal Drug Administration (FDA) in 2022, no data on vaccine effectiveness (VE) of BNT162b2 are available in this age group. We have retrospectively described the safety of BNT162b2 (Pfizer/BioNTech vaccine Comirnaty®) administered off-label in children younger than 5 in years Germany (1). Using data from this authentication-based retrospective survey data obtained between April 14^th^ 2022 and May 9^th^ 2022 (1), we here report VE of BNT162b2 during an Omicron BA.1-2 dominant period.

We analyzed 4615 children aged 2.8±1.2 years (mean ±standard deviation) who received their first dose of BNT162b2 on January 1^st^ 2022 or thereafter (Table S1). We used Cox regression to estimate relative VE of two BNT162b2 doses as indicated in the Supplementary Appendix, with the period between first and second vaccine dose as reference period (24.8±0.6 days) and ≥7 days after Dose 2 to before Dose 3 as Post-vaccination period (59.5±23.6 days).

Table 1 shows that VE was substantial for SARS-CoV-2 infections, symptomatic SARS-CoV-2 infections, and SARS-CoV-2 infections leading to medication use. Differences in dosage of BNT162b2 yielded no change in VE. A sensitivity analysis assessed the geographic differences in VE (Table S2).

**Table 1:** Vaccine effectiveness of two doses of BNT162b2 COVID-19 vaccine during the Omicron BA.1 and BA.2 phase, compared with one dose.

|  | All SARS-CoV-2 infections | Symptomatic SARS-CoV-2 infections | SARS-CoV-2 infections leading to medication use |
| --- | --- | --- | --- |
| **Post-vaccination period** |  |  |  |
| ≥7 Days after Dose 2 to before Dose 3 | 53.1 | 57.5 | 66.2 |
| (p-value) | (<0.001) | (<0.001) | (<0.001) |
| [95%-CI] | [36.3;69.9] | [40.8;74.2] | [43.7;88.7] |
| 3μg | 47.1 | 61.4 | 70.2 |
|  | (<0.001) | (<0.001) | (<0.001) |
|  | [20.7;73.6] | [40.0;82.7] | [44.6;95.7] |
| 5μg | 54.5 | 56.8 | 66.9 |
|  | (<0.001) | (<0.001) | (<0.001) |
|  | [36.6;72.5] | [37.9;75.6] | [43.3;90.5] |
| 10μg | 54.3 | 56.9 | 62.9 |
|  | (<0.001) | (<0.001) | (<0.001) |
|  | [34.9;73.6] | [36.7;77.1] | [32.6;93.2] |
| Infections, n (%) | 779 (16.9) | 621 (13.3) | 261 (5.7) |
| Children, n | 4615 | 4615 | 4615 |

The vaccine effectiveness in % is estimated by $\widehat{VE} = 100 \times (1 - \exp\{\hat{\beta}_{PVP}\})$ , , where $\hat{\beta}_{PVP}$ is the estimated coefficient for the Post-vaccination period of a Cox model stratified by region specific (north, west, east, south, and abroad) calendar day. As control variables, medication use, prior chronic diseases, age, sex, weight, and dosage information of the first vaccination are included.

The present analysis showed that in comparison to one dose of BNT162b2 alone, children receiving a second dose of BNT162b2 had a substantially lower risk for being diagnosed with a SARS-CoV-2 infection or experiencing a SARS-CoV-2 infection leading to symptoms or medication use. The current data contain some limitations. First, children are rarely tested for SARS-CoV-2 and often do not seek medical attention for SARS-CoV-2 symptoms. However, this study coincided with a time when mandatory school/institution testing for SARS-CoV-2 was common in Germany. Next, the assessed vaccination strategy of BNT162b2 was not the one approved by EMA and FDA, with two instead of three BNT162b2 but higher dosages than 3μg in most participants. Furthermore, the reported VE does not necessarily correspond to the currently circulating SARS-CoV-2 variants. Finally, the present data are retrospective and await confirmation by prospective and randomized studies. In conclusion, this study offers a first industry-independent insight in the potential VE of the BNT162b2 vaccine in children aged below 5 years at a time when only immunogenicity data by the manufacturer Pfizer/BioNTech are available (2).

## Data Availability

The raw data are available from the corresponding author on request, provided that a positive evaluation by the pertinent ethics committee is available and German data protection laws are followed.

## Notes

### Competing Interest Statement

OR is supported by the Sara Borrell Fellowship from the Instituto de Salud Carlos III (CD19/00110). JM received funding from the Gottfried & Julia Bangerter-Rhyner Foundation. NT and RB were supported by the coverCHILD project of the German Network University Medicine. RB was also supported by the Federal Ministry of Education and Research (BMBF), Germany, and the Saxon State Ministry for Science and the Arts (SMWK), Federal State of Saxony, Germany. CMC was supported by University Medical Center Rostock. MBM is supported by the Swiss National Science Foundation (grant no. 214187).

### Author Declarations

The Ethics Committee of University of Rostock, Germany gave ethical approval for this work (study ID: A2022-0065).

