## Supplementary appendix for "Vaccine effectiveness of BNT162b2 mRNA Covid-19 Vaccine in Children below 5 Years in German Primary Care"

Data availability ……………………………………………………….	p.6

Figure S1 ……………………………………………………………….	p.7

Table S1 ………………………………………………………………..	p.8

Table S2 ………………………………………………………………..	p.9

Table S3 ………………………………………………………………..	p.10

Table S4 ………………………………………………………………..	p.11

Supplementary references ………………………………………………	p.13

**Supplementary methods**

*Population and vaccinations:*

Design of the authentication-based CoVacU5 survey, recruitment of participants and the authentication procedure have previously been described (1). The CoVacU5 study protocol was approved by the Ethics Committee of University of Rostock, Germany (study ID: A2022-0065). The study was registered in the German Clinical Trials Register (ID: DRKS00028759) and was conducted adhering to the Declaration of Helsinki. The authors vouch for the fidelity to the approved study protocol. Legal guardians of the studied children gave informed consent to participate in the CoVacU5 study. The vaccines themselves were administered independently from the CoVacU5 study. Accordingly, the legal guardians of the studied children had previously given informed consent to the vaccinations. The physicians responsible for the vaccinations had informed them about potential side effects and about the liability of off-label medicine use under the German law. Independently from this study, the physicians are also required to report all unexpected or severe side effects experienced by the children to the German authorities.

In the present analysis, we used the authenticated participants of the CoVacU5 dataset (1) and excluded those observations with vaccinations prior to January 1st 2022 (n=2045) and with prior SARS-CoV-2 infections (n=113). Further, n=331 datasets with missing information about the vaccination date and n=26 datasets with missing information about the weight were excluded from the analysis resulting in a sample of n=4615 observations (Figure S1, Table S1).

*Definition of follow-up periods*

We considered the observation days of the period between first and 1 day before the second BNT162b2 administration as reference, as a proxy for unvaccinated individuals (2,3). We followed the children up to the first positive PCR or antigen-test for SARS-CoV-2 or until 09^th^ of May 2022, whichever came first and censored individuals upon a receipt of third dose of BNT162b2 vaccine. We defined all observation days from day 7 after the second BNT162b2 administration until the day before receiving a third dose of BNT162b2 as the “vaccinated follow-up” period.

*Outcomes*

Co-primary outcomes of the study were all SARS-CoV-2 infections, symptomatic SARS-CoV-2 infections, or SARS-CoV-2 infections leading to medication use. Secondary outcomes were the different dosage strata of BNT162b2 and the sex-stratified analysis of vaccination effectiveness (VE).

*Statistical analysis*

All eligible participants were included in the present analysis without sample size estimations. Statistical analysis was performed using STATA version 15 according to the statistical analysis plan available in the supplementary appendix. VE was estimated as 100*(1-HR) using Cox regression models. VE was calculated as $\hat{VE}=100\times(1-\exp\left\{ \hat{\beta}_{PVP} \right\}$), where $\hat{\beta}_{PVP}$ is the estimated coefficient for the Post-vaccination period of a Cox model stratified by region specific calendar day. This region stratification was chosen to control for potentially different baseline SARS-CoV-2 infection rates over time, and it included 5 regions (northern, western, eastern, southern region of Germany, and abroad). Confounders included in the models were medication use, prior chronic diseases, age, sex, weight. Further, we included dummy variables to control for the more precise geolocation on the federal state level. Finally, since the population included children vaccinated with different doses of BNT162b2, we investigated the effects of dosage in an interaction term together with the follow-up period. Two-tailed p values of <0.05 were considered statistically significant. A sensitivity analysis was performed with stratification for geolocalization in 5 large regions as reported in Table S2. A sex-stratified analysis and the hazards of all included model parameters are shown in Tables S2 and S3. Table S4 shows the hazard of each of the included model parameters. Finally, an additional robustness check of the data was performed by multiple imputation of the weight and vaccination dosage in the dataset of 4979 children (data not shown). No adjustments were made for multiplicity.

**Author Contributions:**

Dr Strumann had full access to all of the data in the study and takes responsibility for the integrity of the data and the accuracy of the data analysis. Drs M.B. Moor and Chao are co–senior authors.

*Concept and design:* Strumann, Ranzani, J. Moor, Chao, M.B. Moor.

*Acquisition, analysis, or interpretation of data:* Strumann, Ranzani, J. Moor, Berner, Toepfner, Chao, M.B. Moor.

*Drafting of the manuscript:*  M.B. Moor.

*Critical revision of the manuscript for important intellectual content:* Strumann, Ranzani, J. Moor, Berner, Toepfner, Chao, M.B. Moor.

*Statistical analysis:* Strumann.

*Administrative, technical, or material support:* Toepfner, Berner, Chao.

*Supervision:* Chao, Moor.

**Conflict of Interest Disclosures:**

OR is supported by the Sara Borrell Fellowship from the Instituto de Salud Carlos III (CD19/00110). JM received funding from the Gottfried & Julia Bangerter-Rhyner Foundation. NT and RB were supported by the coverCHILD project of the German Network University Medicine. RB was also supported by the Federal Ministry of Education and Research (BMBF), Germany, and the Saxon State Ministry for Science and the Arts (SMWK), Federal State of Saxony, Germany. CMC was supported by University Medical Center Rostock. MBM is supported by the Swiss National Science Foundation (grant no. 214187). No other disclosures were reported.

**Acknowledgements:**

The authors wish to thank all participating vaccination centers and respondents. The authors acknowledge the contributions to database design by Denisa Drinka, MD and Maximilian Jorczyk, MD (Technische Universität Dresden, Germany) and for participant recruitment by Wolfgang von Meißner, MD (Hausärzte am Spritzenhaus, Baiersbronn, Germany), David Stuppe, MD (University Medical Center, Rostock, Germany), Johannes Püschel, MD (Hausarztzentrum, Greven, Germany), Melanie Liss, MD (Praxis für Kinder- und Jugendmedizin, Düsseldorf, Germany), Emilie von Poblotzki (Praxis die Kinderärzte, München, Germany), Anke Böhnke, MD (Frauenarztpraxis, Berlin, Germany), Armin Philipp, MD (Hautarztpraxis, Stuttgart, Germany), Georg Hillebrand, MD (Klinik für Kinder- und Jugendmedizin, Itzehoe, Germany), Lisa Degener, MD (Hausärztliche Gemeinschaftspraxis, Altenberge, Germany), and Lisa Schneider, MD (Gemeinschaftspraxis Dres Albrecht/Winkler, Laupheim, Germany). We acknowledge the nonprofit assistance for participant recruitment provided by the Bildung Aber Sicher and U12Schutz initiatives. None were compensated for their work on the CoVacU5 study.

**Data availability**

The raw data are available from the corresponding author on request, provided that a positive evaluation by the pertinent ethics committee is available and German data protection laws are followed.

**Supplementary figure**

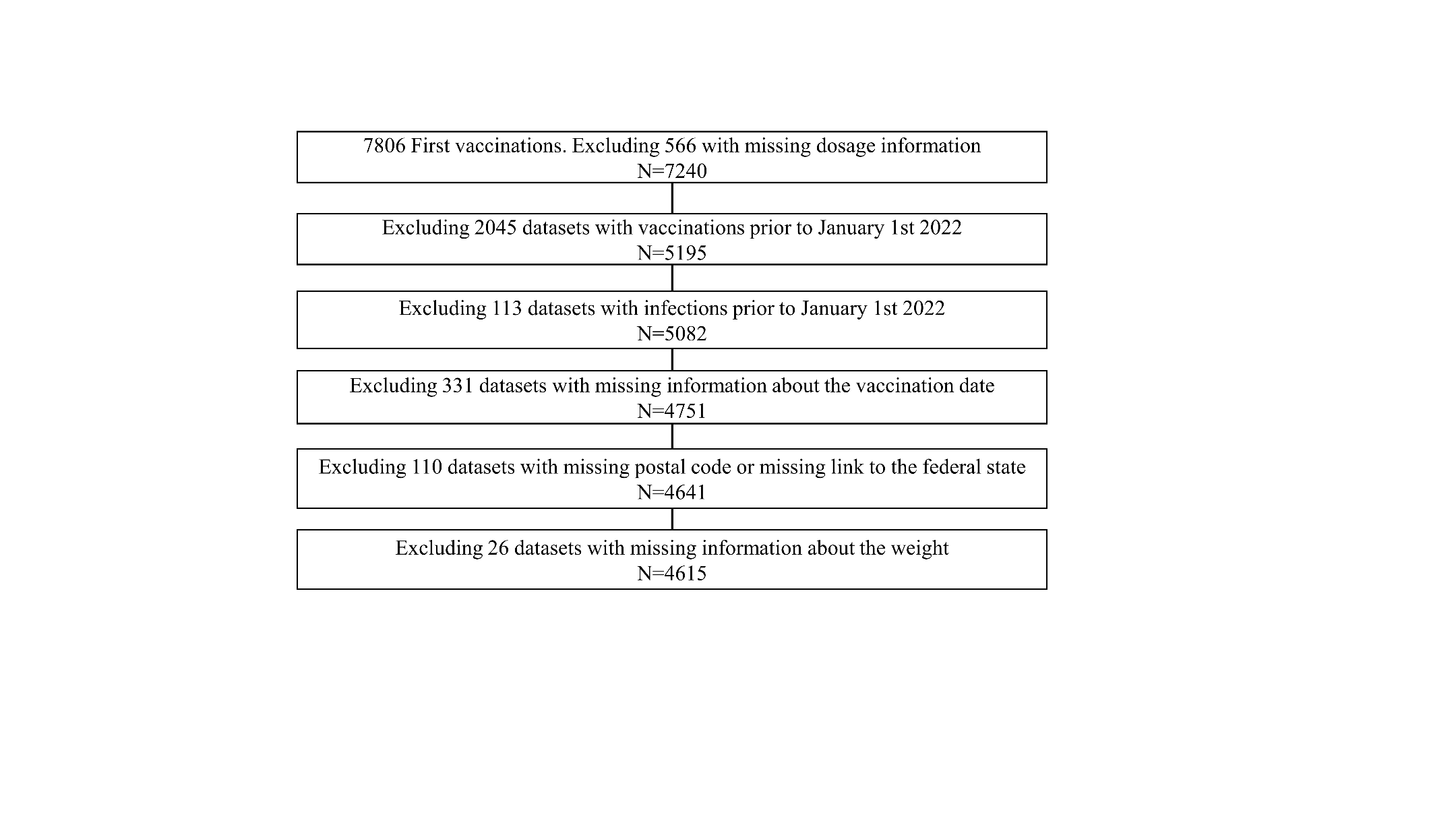

Figure S1. Flow-chart of study participants.

**Supplementary tables**

**Table S1. Characteristics of Children**

| **Characteristic** | **Total** | **analyzed sample** |
| --- | --- | --- |
|  | **n=4979** | **n=4615** |
| ***Sex*** |  |  |
| Female | 2439 (49.0) | 2262 (49.0) |
| Male | 2539 (51.0) | 2352 (51.0) |
| Divers | 1 (0.0) | 1 (0.0) |
| Age, median (IQR), y | 3 (2-4) | 3 (2-4) |
| Weight, median (IQR), kg | 14 (12-17) | 14 (12-16.5) |
| ***Dosage at first vaccination, μg*** |  |  |
| 3 | 1772 (24.5) | 833 (18.0) |
| 5 | 3120 (43.1) | 2162 (46.8) |
| 10 | 2348 (32.4) | 1620 (35.1) |
| Long-term medication | 239 (4.8) | 217 (4.7) |
| Comorbidities (yes) | 409 (8.2) | 376 (8.1) |
| ***Regional variables*** |  |  |
| Abroad | 267 (5.4) | 243 (5.3) |
| Schleswig Holstein | 276 (5.5) | 234 (5.1) |
| Hamburg | 201 (4.0) | 180 (3.9) |
| Lower Saxony | 562 (11.3) | 539 (11.7) |
| Bremen | 50 (1.0) | 49 (1.1) |
| North Rhine-Westphalia | 1194 (24.0) | 1085 (23.5) |
| Hesse | 313 (6.3) | 281 (6.1) |
| Rhineland Palatinate | 269 (5.4) | 256 (5.5) |
| Baden-Württemberg | 790 (15.9) | 766 (16.6) |
| Bavaria | 649 (13.0) | 591 (12.8) |
| Saarland | 35 (0.7) | 32 (0.7) |
| Berlin | 343 (6.9) | 329 (7.1) |
| Brandenburg | 96 (1.9) | 93 (2.0) |
| Mecklenburg Western Pomerania | 33 (0.7) | 31 (0.7) |
| Saxony | 108 (2.2) | 101 (2.2) |
| Saxony-Anhalt | 32 (0.6) | 29 (0.6) |
| Thuringia | 41 (0.8) | 38 (0.8) |

The analyzed sample was used for the main analyses. The total population included

missing data in weight and/or vaccine doasage and was used for multiple imputation.

Brackets indicate % unless stated otherwise.

**Table S2: Vaccination effectiveness of BNT162b2 separated per region**

| **region** | **All SARS-CoV-2 infections**  % (p-value)  [95%-CI] | **Symptomatic SARS-CoV-2 infections**  % (p-value)  [95%-CI] | **SARS-CoV-2 infections leading to medication use**  % (p-value)  [95%-CI] |
| --- | --- | --- | --- |
| **Northern** | 35.6  (0.232)  [-0.227;0.939] | 35.6  (0.268)  [-27.5;98.8] | 56.3  (0.100)  [-10.8;123.3] |
| Infections, n/N (%) | 145/894(16.2) | 119/868(13.7) | 39/788(4.9) |
| **Eastern** | 53.9  (0.005)  [16.4;91.4] | 60.6  (<0.001)  [26.4;94.8] | 86.4  (<0.001)  [65.6;107.2] |
| Infections, n/N (%) | 96/565(17.0) | 77/546(14.1) | 33/501(6.6) |
| **Southern** | 56.1  (<0.001)  [24.3;88.0] | 73.2  (0.000)  [53.7;92.7] | 68.7  (0.025)  [8.8;128.5] |
| Infections, n/N (%) | 228/1319(17.3) | 175/1266(13.8) | 67/1158(5.8) |
| **Western** | 63.0  (<0.001)  [43.5;82.6] | 57.1  (<0.001)  [30.9;83.3] | 61.2  (<0.001)  [23.8;98.5] |
| Infections, n/N (%) | 269/1594(16.9) | 213/1539(13.8) | 98/1423(6.9) |
| **Abroad** | -23.8  (0.893)  [-371.3;323.6] | -3.5  (0.981)  [-294.5;287.5] | 24.3  (0.768)  [-137.0;185.5] |
| Infections, n/N (%) | 42/243(17.3) | 37/238(15.5) | 24/225(10.7) |

N is the number of children used in the estimation; n is the number children that have an infection in the respective region. The post-vaccination period of ≥7 Days after Dose 2 to before Dose 3 was compared with the period between Dose 1 and Dose 2 as a reference. Region refers to residence within Germany.

**Table S3: Sex-stratified analysis of vaccination effectiveness**

|  | **All SARS-CoV-2 infections**  % (p-value)  [95%-CI] | **Symptomatic SARS-CoV-2 infections**  % (p-value)  [95%-CI] | **SARS-CoV-2 infections leading to medication use**  % (p-value)  [95%-CI] |
| --- | --- | --- | --- |
| **females** | 49.0  (<0.001)  [22.7;75.4] | 58.2  (<0.001)  [35.8;80.7] | 55.0  (0.027)  [6.4;103.7] |
| Infections n/N (%) | 354/2262(15.6) | 284/2192(13.0) | 113/2020(5.6) |
| **males** | 54.3  (<0.001)  [31.6;77.0] | 54.0  (<0.001)  [27.0;81.0] | 61.2  (<0.001)  [25.3;97.1] |
| Infections n/N (%) | 425/2352(18.1) | 337/2264(14.9) | 148/2074(7.1) |

N is the number of children used in the estimation; n is the number children that have an infection in the respective region. Post-vaccination period ≥7 Days after Dose 2 to before Dose 3.

**Table S4: Estimated hazard ratios**

| **Variable** | **M1** | **M2** | **M3** | **M4** |
| --- | --- | --- | --- | --- |
| Follow-Up (reference FU1: Dose 1 to before Dose 2) |  |  |  |  |
| FU2: Dose 2 to <7 days after Dose 2 | 0.434*** | 0.429*** | 0.429*** |  |
| FU3: >=7 Days after Dose 2 to before Dose 3 | 0.467*** | 0.464*** | 0.469*** |  |
| ***Dosage specific FU-Effects*** |  |  |  |  |
| FU2 X Dosage 3µg |  |  |  | 0.669 |
| FU2 X Dosage 5µg |  |  |  | 0.383*** |
| FU2 X Dosage 10µg |  |  |  | 0.385*** |
| FU3 X Dosage 3µg |  |  |  | 0.529** |
| FU3 X Dosage 5µg |  |  |  | 0.455*** |
| FU3 X Dosage 10µg |  |  |  | 0.457*** |
| ***Controls*** |  |  |  |  |
| dosage at first vaccination (ref. 3µg) |  |  |  |  |
| 5µg |  | 0.904 | 0.894 | 1.028 |
| 10µg |  | 0.946 | 0.941 | 1.077 |
| age (ref. 0.5) |  |  |  |  |
| 1 |  | 1.165 | 1.168 | 1.163 |
| 1.5 |  | 0.941 | 0.932 | 0.933 |
| 2 |  | 1.591* | 1.588* | 1.579* |
| 2.5 |  | 1.700** | 1.667** | 1.654* |
| 3 |  | 1.966** | 1.952** | 1.937** |
| 3.5 |  | 2.005** | 1.989** | 1.978** |
| 4 |  | 1.976** | 1.982** | 1.970** |
| 4.5 |  | 2.485*** | 2.432*** | 2.414*** |
| female |  | 0.886* | 0.884* | 0.885* |
| weight |  | 0.99 | 0.991 | 0.992 |
| Long-term medication |  | 1.442* | 1.432* | 1.437* |
| Comorbidities (yes) |  | 1.084 | 1.091 | 1.089 |
| ***regional variables*** *(ref. abroad)* |  |  |  |  |
| Schleswig Holstein |  |  | 1.468 | 1.473 |
| Hamburg |  |  | 1.213 | 1.219 |
| Lower Saxony |  |  | 1.296 | 1.299 |
| Bremen |  |  | 0.827 | 0.829 |
| North Rhine-Westphalia |  |  | 1.523 | 1.533 |
| Hesse |  |  | 1.248 | 1.253 |
| Rhineland Palatinate |  |  | 1.326 | 1.336 |
| Baden-Württemberg |  |  | 0.455 | 0.454 |
| Bavaria |  |  | 0.411 | 0.41 |
| Saarland |  |  | 1.892 | 1.899 |
| Berlin |  |  | 1.399 | 1.362 |
| Brandenburg |  |  | 1.601 | 1.571 |
| Mecklenburg Western Pomerania |  |  | 2.105** | 2.069** |
| Saxony |  |  | 1.284 | 1.241 |
| Saxony-Anhalt |  |  | 1.853 | 1.779 |
| Thuringia |  |  | 1.673 | 1.601 |
| LOGLIKE | -2372.0 | -2341.3 | -2334.8 | -2333.8 |
| AIC | 4748.1 | 4714.6 | 4733.7 | 4739.6 |
| BIC | 4769.8 | 4888.6 | 5081.6 | 5131.1 |

* p<.1; **p<.05; *** p<.01; observations from 4615 children used in the estimation; stratified by region specific (north, west, east, south, and abroad) calendar day; LOGLIKE: Loglikelihood value; AIC: Akaike information criterion; BIC: Bayesian information criterion
